# Association between Weather Variables and Viral Gastroenteritis in the United States

**DOI:** 10.64898/2026.04.29.26352095

**Authors:** Natalya Alekhina, Paola Fonseca-Romero, Per H. Gesteland, Ben J. Brintz, Amy L. Leber, Jami T. Jackson, Jennifer Dien Bard, Neena Kanwar, Ara Festekjian, Chari Larsen, Kimberle C. Chapin, Rangaraj Selvarangan, Sean Soisson, Andrew T. Pavia, Daniel T. Leung

## Abstract

Infectious gastroenteritis (IGE) is a major cause of pediatric morbidity globally, with viral pathogens accounting for a substantial proportion of cases. While seasonal patterns of viral IGE are well recognized, the association between specific environmental exposures, such as ambient temperature, and viral IGE has not been fully quantified.

First, we performed a secondary analysis of data from a prospective, multisite study of children presenting to emergency departments at five medical centers across the continental United States, linking individual level laboratory data to environmental exposures, including temperature, humidity, and air pollutants, measured during the 14 days preceding symptom onset. Mixed-effects logistic regression was applied to evaluate the association between viral IGE and environmental exposures, adjusting for site-level clustering and patient age. Among 868 children with IGE, higher ambient temperature was inversely associated with viral etiology (OR 0.50, 95% CI 0.36–0.68, p < 0.001). We did not find statistically significant associations between other environmental variables and viral IGE.

Then, to contextualize these individual-level findings in children, we examined all-ages population-level surveillance data from GermWatch, a regional laboratory testing-based infectious disease surveillance system, which demonstrated concordant declines in viral pathogen detection with increasing temperature.

These findings support the association of weather with viral transmission patterns. Incorporating environmental context into clinical decision-making may improve diagnostic stewardship and support more effective resource allocation during periods of increased viral IGE prevalence.

## INTRODUCTION

Infectious gastroenteritis (IGE) remains a significant cause of pediatric morbidity and mortality in both high- and low-income countries. While laboratory diagnostic testing informs management decisions, excessive testing increases healthcare costs and may lead to inappropriate antimicrobial use.^1^ Understanding how external factors, such as weather variables, relate to IGE etiology is therefore clinically relevant as it can help clinicians better interpret the seasonality of IGE and guide appropriate use of diagnostics.^2^

Evidence indicates that temperature, humidity, and precipitation influence infectious disease transmission.^3,4^ Weather variables have also been incorporated into clinical prediction models in other settings, where they improved discriminative performance,^5–7^ underscoring the broader value of understanding these relationships. At the population level, weather patterns can inform expectations for pathogen circulation and help guide public health preparedness.

Although prior studies have shown that viral IGE, particularly norovirus and rotavirus, follows a seasonal pattern in the US, with peaks during the winter months,^8^ the association of temperature and other environmental factors on the overall incidence of viral gastroenteritis has not been well quantified. To better understand the seasonality of viral IGE, we assessed the association between environmental factors and viral IGE in a prospective multicenter study of children presenting to emergency departments with acute gastroenteritis and confirmed our findings using all-ages population-level surveillance data from a regional infectious disease monitoring system.

## METHODS

### Dataset and Cohort Construction

We first utilized data from the Implementation of a Molecular Diagnostic for Pediatric Acute Gastroenteritis (IMPACT) study,^9^ a prospective pragmatic stepped wedge trial evaluating the clinical impact of multiplex polymerase chain reaction (PCR) testing for children under 18 years presenting with gastroenteritis.

The dataset included 1,157 pediatric patients who presented with acute IGE to emergency departments (EDs) at five major medical centers in the continental United States between March 26, 2015, and August 22, 2016. For this analysis we included all enrolled subjects who provided a stool specimen for laboratory evaluation. Stool pathogens were identified using the BioFire FilmArray GI Panel, a multiplex PCR assay^9^, with few clinician-ordered viral detections confirmed by targeted PCR. We excluded patients with completely missing pathogen results (as opposed to not detected) from etiological analysis. EPEC (Enteropathogenic *Escherichia coli*) detections (in patients of any age) were re-classified as “not detected” due to the high prevalence of asymptomatic carriage in the general population.^10^ Likewise, *C. difficile*^9^ detections in children younger than two years were reclassified as “not detected” as *C. difficile* colonization is very common in this age group.^11^

Detected pathogens were first classified as viral, bacterial, or protozoal, and stool samples containing multiple organisms were labeled according to all pathogen types present. To align with the study’s focus on viral IGE, we categorized samples containing only viral pathogens as “viral,” while samples containing only bacterial pathogens or a mixture of pathogen types were categorized as “other.” Samples without a detected pathogen (n = 298) were classified as “unidentified” (**Table 1**) and excluded from the primary analysis to minimize misclassification bias.

**Table 1.** Patient Distribution by Infectious Gastroenteritis Etiology (IMPACT)

| IMPACT Dataset |  |  |  |
| --- | --- | --- | --- |
| Original Pathogen Classification |  | Final Pathogen Classification |  |
| Pathogen Category | Patients (n=868) | Pathogen Category | Patients (n=868) |
| Virus | 337 (38.8%) | Viral | 337 (38.8%) |
| Bacteria | 138 (15.9%) | Other | 233 (26.8%) |
| Bacteria + Virus | 56 (6.5%) |  |  |
| Protozoa | 22 (2.5%) |  |  |
| Protozoa + Bacteria | 8 (0.9%) |  |  |
| Protozoa + Virus | 7 (0.8%) |  |  |
| Protozoa + Bacteria + Virus | 2 (0.2%) |  |  |
| Unidentified/None | 298 (34.3%) | Unidentified | 298 (34.3%) |

### Environmental Data

We obtained environmental data from publicly available sources: daily temperature and precipitation from the National Oceanic and Atmospheric Administration (NOAA),^12^ daily relative humidity from the Historical Weather API (Open Meteo),^13^ and air pollution data (carbon monoxide, lead, nitrogen dioxide, and ozone) from the United States Environmental Protection Agency (EPA)^14^ at each of the five sites. For each clinical site, we extracted 14 days of environmental data per patient, covering the date of IGE onset and the 13 preceding days. To account for variation in proximity between observation stations and clinical sites, we used weighted estimates by calculating the distance between each station and the corresponding ED location with the Haversine formula.^15^ We computed daily weighted averages for temperature, precipitation, humidity, and air pollutants using the inverse distance weighting based on each station’s proximity to the clinical site. Across pollutants (CO, Pb, NO□, O□), the proportion of missing daily observations ranged from <0.40% to 8.21%. We imputed missing pollutant values in a stepwise site-specific approach. First, we replaced missing daily observations with monthly means for the same site, calendar month, and year. If no observations were available for that month, we used the mean from the preceding month, followed by the subsequent month, and finally the overall site mean. This procedure eliminated all missing pollutant values.

### Individual-Level Modeling (IMPACT Dataset)

We calculated a 14-day mean for each environmental variable per patient, condensing the data to one row per individual. These 14-day averages served as predictors, with IGE etiology (viral versus “other”) as the binary outcome. We used mixed effects logistic regression to account for within-site correlation. All continuous exposure variables were standardized (mean = 0, SD = 1) to facilitate model convergence and allow for comparison across predictors. Age was included as a covariate to account for age-related variability in the incidence of viral IGE and was modeled using the following age groups (<2 years; ≥2 to <5 years; ≥5 to <12 years; ≥12 years). **Supplemental Table 1** captures viral IGE distribution across age groups. To minimize misclassification, we excluded 298 patients with unidentified IGE etiology from the final analytic sample.

### Population-Level Modeling (GermWatch Dataset)

To confirm our individual-level findings at the population level, we evaluated the association between viral IGE activity and environmental factors using data from GermWatch,^16^ an all-ages regional infectious disease surveillance system that aggregates results of routine stool microbiologic testing from clinics, emergency departments and hospitals across Utah. GermWatch compiles laboratory results generated through clinician-directed diagnostic testing, providing a complementary real-world perspective to the protocol-driven sampling used in the IMPACT study. For comparability with the multiplex PCR methodology used in IMPACT, we restricted analyses to January 2015 through August 2025, when multiplex polymerase chain reaction (PCR) stool panels, including the BioFire FilmArray GI Panel, became broadly available within the facilities of the surveillance system and was the most common methodology used. The dataset included monthly percentages of individual pathogens detected in stool samples, with analysis further restricted to two counties Utah and Salt Lake counties, counties with the highest proportion of testing and for which geographic location could be reliably specified (**Table 2**). This allowed linkage of pathogen surveillance data with environmental exposures using a county-level centroid-based approach. For each county, we identified geographic centroids and selected the nearest monitoring stations with the most complete reporting. This centroid-based nearest-station approach is consistent with prior Utah studies on county-level exposure assessment.^17^ Using the same environmental data sources applied in the IMPACT analysis, we extracted monthly estimates of mean temperature, relative humidity, and ambient carbon monoxide concentrations for each county.

**Table 2.** County-Level Distribution of Pathogens (GermWatch)

| GermWatch Dataset |  |  |  |  |
| --- | --- | --- | --- | --- |
| Final Pathogen Classification | County | Mean Percent Positive | Standard Deviation | Number of Months |
| Viral | Salt Lake | 3.8% | 4.0% | 622 |
| Viral | Utah | 4.5% | 7.2% | 604 |
| Other | Salt Lake | 1.4% | 1.4% | 1018 |
| Other | Utah | 1.5% | 2.5% | 1004 |

After classifying pathogens as viral or “other”, we calculated the monthly proportion of viral detections for each county and used an autoregressive (ARIMAX) model to evaluate the relationship between the current month’s viral proportion and prior-month viral incidence, while accounting for concurrent environmental exposures. The monthly time-series structure of the GermWatch dataset warranted use of an ARIMAX framework to account for temporal dependence and incorporate environmental covariates. ARIMAX models were fitted using the auto.arima() function (forecast package in R) with a seasonal period of 12 months. The function selected the best-fitting model by minimizing the corrected Akaike Information Criterion and ensuring an appropriate balance between model fit and parsimony. This approach was well suited for infectious disease surveillance data, which typically exhibit temporal autocorrelation and strong seasonal structure, and allows estimation of weather effects after adjusting for these inherent temporal patterns. With the outcome expressed as a continuous monthly proportion and only infrequent values near zero, the data met the assumptions needed to specify the ARIMAX models with a linear (identity) link, predicting viral proportion on its original scale. Accordingly, ARIMAX coefficients were interpreted as linear effects rather than odds ratios.

## PATIENT CONSENT STATEMENT

This secondary data analysis was reviewed by the IRB of the University of Utah and determined to be exempt (IRB_00166050).

## RESULTS

Among 1,157 children presenting with IGE in the IMPACT cohort, 868 submitted a stool sample and were included in etiologic analyses (**Table 1**). Viral-only etiologies accounted for 337 cases (38.8%), while 233 cases (26.84%) were classified as “other” (non-viral or mixed etiologies), and 298 cases (34.3%) had no pathogen detected. Children younger than two years accounted for the majority of viral IGE cases (57.3%), highlighting the higher prevalence of viral IGE in this age group.

We found a statistically significant association between temperature and viral IGE etiology using the IMPACT dataset (**Table 3**). Specifically, a 1 standard deviation increase in average temperature in the prior 14 days was associated with a 50% reduction in the odds of viral etiology (OR = 0.50, 95% CI: 0.36-0.68, p < 0.001), after adjustment for other environmental variables and age. Compared with younger than two years, progressively lower odds of viral IGE were observed in older age groups, with adolescents (≥12 years) demonstrating 83% lower odds of viral IGE (OR = 0.17, 95% CI: 0.09-0.33, p < 0.001), after adjusting for environmental exposures and site. Other environmental exposures were not statistically significant predictors of viral IGE (p > 0.05). We also visually demonstrated the relationship between calendar month, ambient temperature, and viral IGE prevalence (**Supplemental Figure 1**), and the direct relationship between ambient temperature and IGE prevalence (**Supplemental Figure 2)**.

**Table 3.**
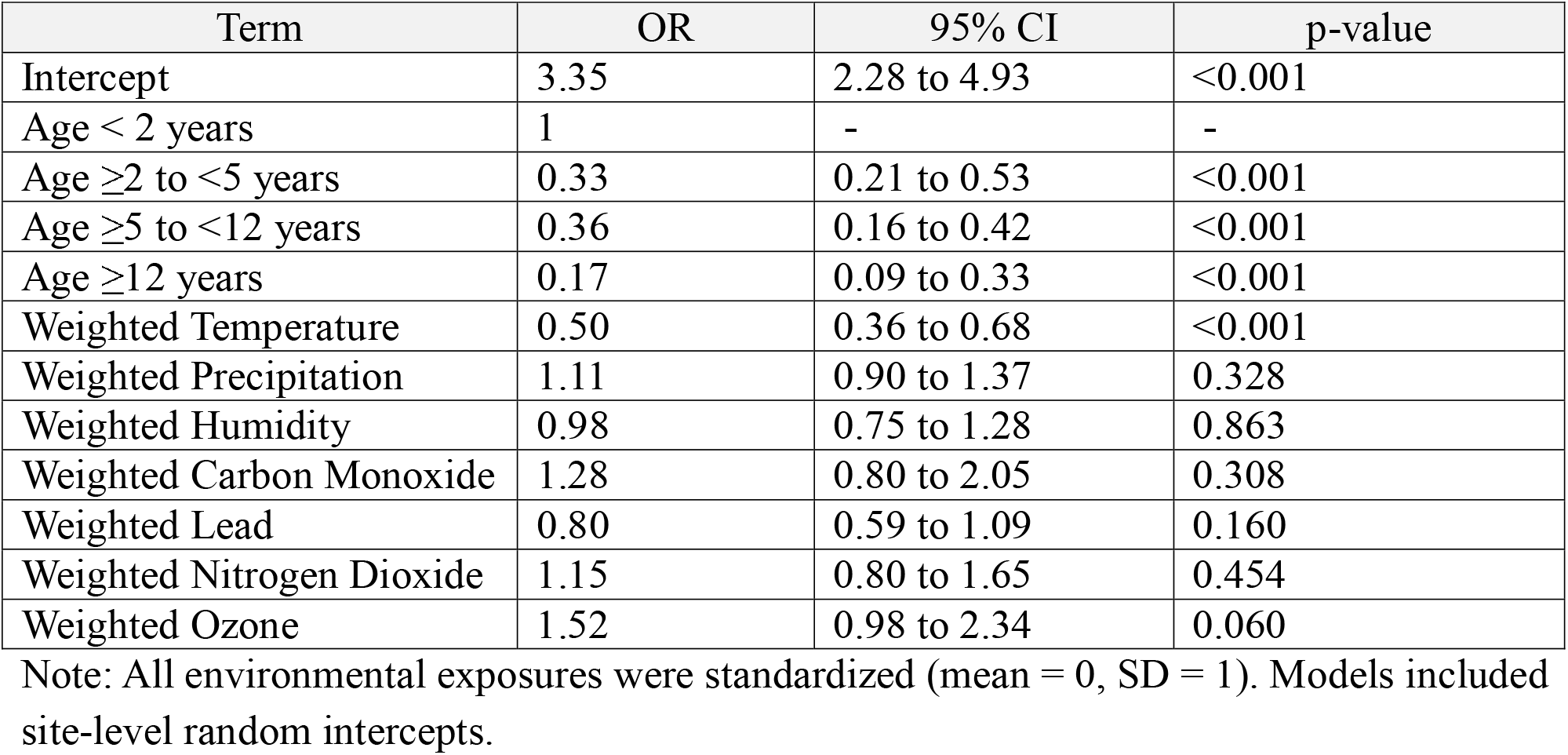
Mixed Effects Logistic Regression of Viral vs. Non-Viral Gastroenteritis (IMPACT)

| Term | OR | 95% CI | p-value |
| --- | --- | --- | --- |
| Intercept | 3.35 | 2.28 to 4.93 | <0.001 |
| Age < 2 years | 1 | - | - |
| Age ≥2 to <5 years | 0.33 | 0.21 to 0.53 | <0.001 |
| Age ≥5 to <12 years | 0.36 | 0.16 to 0.42 | <0.001 |
| Age ≥12 years | 0.17 | 0.09 to 0.33 | <0.001 |
| Weighted Temperature | 0.50 | 0.36 to 0.68 | <0.001 |
| Weighted Precipitation | 1.11 | 0.90 to 1.37 | 0.328 |
| Weighted Humidity | 0.98 | 0.75 to 1.28 | 0.863 |
| Weighted Carbon Monoxide | 1.28 | 0.80 to 2.05 | 0.308 |
| Weighted Lead | 0.80 | 0.59 to 1.09 | 0.160 |
| Weighted Nitrogen Dioxide | 1.15 | 0.80 to 1.65 | 0.454 |
| Weighted Ozone | 1.52 | 0.98 to 2.34 | 0.060 |

Note: All environmental exposures were standardized (mean = 0, SD = 1). Models included site-level random intercepts.

To assess multicollinearity among environmental predictors, we calculated variance inflation factors (VIFs) using an equivalent fixed-effects logistic regression model. All adjusted VIF values were below 2, indicating that multicollinearity was not a concern.

As a sensitivity analysis, we repeated the model after reclassifying cases of unidentified etiology as “other” (**Supplemental Table 2**). The direction and magnitude of primary associations were largely similar, with higher temperature (OR = 0.61, CI: 0.50 to 0.76, p < 0.001) and older age (OR = 0.24, CI: 0.14 to 0.42, p < 0.001) remaining significantly associated with lower odds of viral IGE. Higher humidity was associated with lower odds of viral IGE (OR = 0.8, CI: 0.66 to 0.97, p = 0.026), while higher carbon monoxide levels were associated with increased odds (OR = 1.29, CI: 1.06 to 1.58, p = 0.012) in this sensitivity analysis but not in the primary analysis.

Consistent with these individual-level findings in children, population-level GermWatch surveillance data featuring all ages demonstrated an inverse association between prior-month ambient temperature and the proportion of detected viral pathogens (**Figure 1**). At the county level, ARIMAX indicated that lower mean monthly temperatures were significantly associated with a higher proportion of viral gastroenteritis cases in both Salt Lake County (β = −0.0028 per °F; 95% CI, −0.0048 to −0.0008; p = 0.007) and Utah County (β = −0.0063 per °F; p = 0.007) (**Supplemental Figure 3**). Relative humidity was modestly inversely associated with the proportion of viral cases in Salt Lake County (β = −0.0019; p = 0.04) but not in Utah County. Carbon monoxide showed no significant association in either model. Analysis of data from both counties were combined yielding consistent results: mean temperature remained a significant inverse predictor of proportion of viral cases (β = −0.0052 ± 0.0017 per °F; p = 0.002), while humidity and CO were not significant (**Table 4**). All ARIMAX models demonstrated good fit (RMSE range: 0.09–0.23) and no residual autocorrelation (Ljung–Box p ≥ 0.36), with fitted values closely tracking observed seasonal peaks in viral activity during cooler months (**Supplemental Figure 4 and Supplemental Figure 5**).

**Figure 1.**
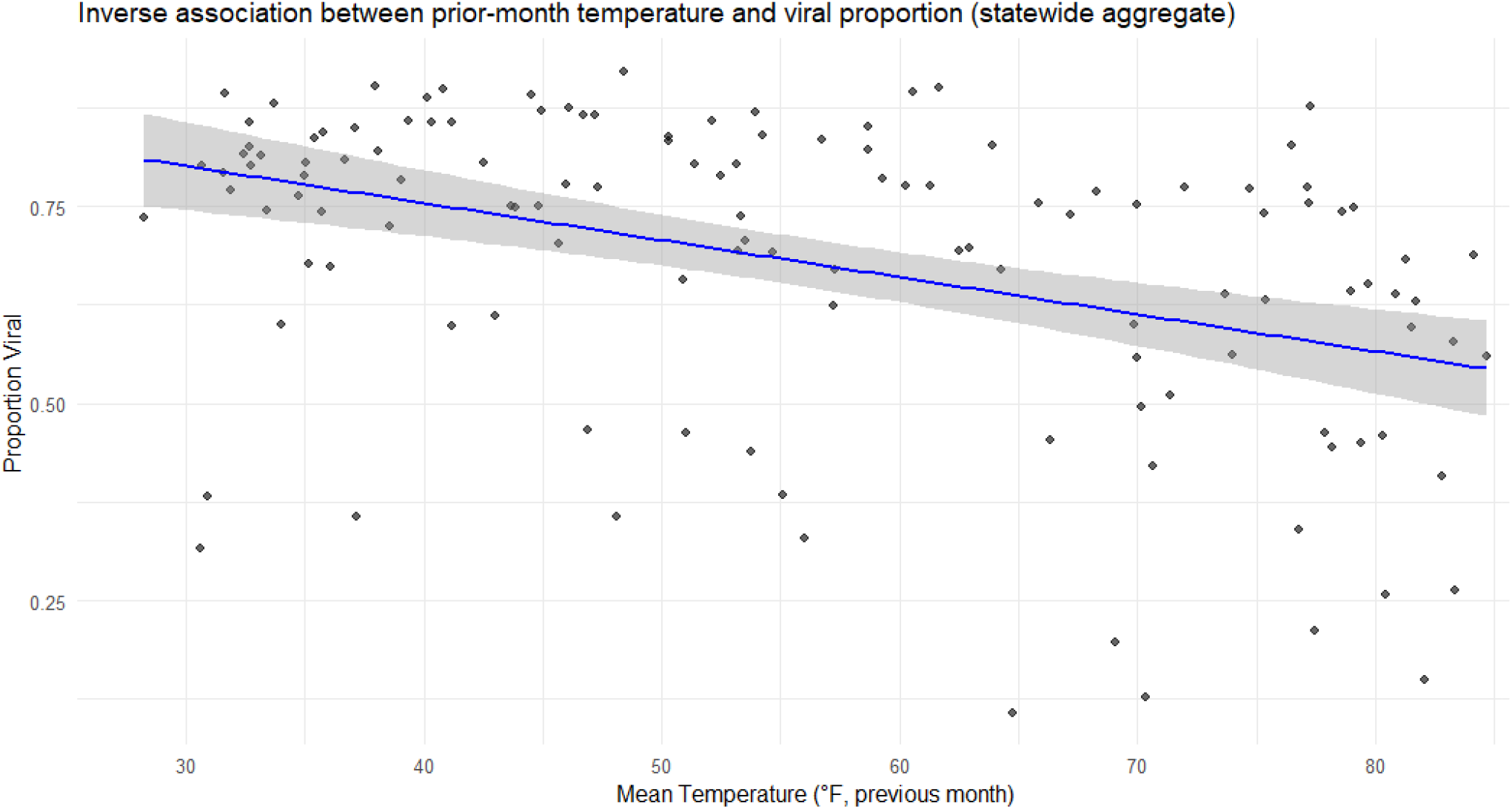
Prior Month Temperature vs. Proportion of Viral Pathogens - Aggregated (GermWatch) Monthly mean ambient temperature from the preceding month was plotted against the proportion of stool test detections attributed to viral pathogens using statewide aggregated GermWatch surveillance data. Each point represents a single month, with the proportion calculated as the number of viral pathogen detections divided by the total number of identified pathogens for that month. The solid line represents the fitted values from an ordinary least squares linear regression model, with the shaded area indicating the 95% confidence interval, illustrating the overall inverse association between prior-month temperature and viral pathogen proportion.

**Table 4.** Autoregressive Integrated Moving-Average (ARIMAX) Model Output (GermWatch)

| County | Term | $\beta$ (per °F) | SE | 95% CI | p-value | ARIMA Model |
| --- | --- | --- | --- | --- | --- | --- |
| Salt Lake | Mean temperature (°F) | -0.0028 | 0.0010 | -0.0048 to 0.0008 | 0.007 | ARIMA(1,0,0)(0,1,1)[12] |
| Salt Lake | Relative humidity (%) | -0.0019 | 0.0009 | -0.0037 to 0.0001 | 0.042 |  |
| Utah | Mean temperature (°F) | -0.0063 | 0.0023 | -0.0108 to 0.0018 | 0.007 | ARIMA(0,0,1)(0,1,1)[12] |
| Aggregate | Mean temperature (°F) | -0.0052 | 0.0017 | -0.0085 to 0.0019 | 0.002 | ARIMA(1,0,1)(0,1,1)[12] |

## DISCUSSION

Our analysis demonstrated a significant inverse association between average daily temperature and viral IGE, refining the characterization of known seasonal patterns. Our findings are consistent with studies in Asia involving substantially different climates,^18–21^ and held true for both individual-level and population-level datasets analyzed. In the pediatric cohort, older age was independently associated with lower odds of viral IGE, reflecting well-established epidemiologic patterns. Importantly, inclusion of age as a covariate did not materially attenuate the observed association between ambient temperature and viral etiology, supporting the robustness of the relationship between temperature and viral IGE.

Our findings have implications at both individual and population levels. At the individual level, incorporating weather variables into clinical prediction tools may help prioritize diagnostic testing, minimize unnecessary antibiotic use, and improve the timely diagnosis and management of IGE. At the population level, understanding the impact of temperature on the incidence of viral IGE can inform more strategic resource allocation, particularly in settings with limited access to PCR stool testing. Because GermWatch reflects real-world clinician testing behavior within a large integrated delivery system, these findings demonstrate how population-level surveillance data can complement individual-level clinical prediction efforts. Integrating such regional epidemiologic signals into diagnostic clinical decision support may help clinicians calibrate pre-test probability for viral etiologies and avoid unnecessary stool testing during predictable seasonal peaks. Moreover, weather-informed surveillance models can enhance the accuracy of seasonal outbreak predictions and optimize healthcare delivery during seasonal surges.

As global temperatures rise, seasonal patterns of infectious diseases, including gastroenteritis, are likely to shift, underscoring the importance of continued monitoring and adaptable modeling strategies. Our findings provide a foundation for developing weather-informed models to support more scalable healthcare resource allocation in the context of climate change. As weather patterns vary year to year and with changing climate, weather informed models are likely to prove superior to those based on season.

This study has several limitations. First, the analysis was restricted to data collected within the continental United States, which may limit the global generalizability of our findings. Second, the relatively short duration of available individual-level data may have limited our ability to capture long-term seasonal patterns or rare events. Third, several air pollution variables had substantial missingness, requiring imputation and potentially affecting the robustness of the exposure estimates. Fourth, site-level sample sizes were small at several clinical locations, reducing the precision of site-specific estimates and limiting power to detect consistent associations across sites. Fifth, the GermWatch dataset reflected testing ordered at clinicians’ discretion, introducing potential selection bias – a known limitation of surveillance systems based on routine diagnostic workflows rather than systematic sampling. Finally, environmental exposures were assigned based on conditions near the emergency department rather than patients’ residential locations, which were unavailable. Although weather patterns are generally consistent across the catchment areas served by participating sites, air pollutant concentrations may vary over larger geographic regions, potentially introducing exposure misclassification.

Future studies should focus on collecting more comprehensive and longitudinal datasets and on obtaining more complete air pollution data and patient-level location information to improve exposure assignment. Additionally, similar analysis should be conducted in diverse geographic and socioeconomic contexts outside of United States. Nevertheless, we found a significant inverse association between average daily temperature and the odds of viral IGE. These findings support the utility of incorporating weather variables, particularly temperature, into clinical prediction models to improve the identification of viral IGE. Future models leveraging real-time geographically diverse environmental data may help guide clinical decision-making, especially in settings where diagnostic resources are limited.

## Data Availability Statement

Data are available upon request. Analytic code is available in GitHub pending final peer review.

## Author Contributions

N.A. conceptualized the study, performed data curation and statistical analysis, interpreted the results, and drafted the manuscript

P.F.R. contributed to data curation and analysis and revised the manuscript.

P.G.H. contributed to data acquisition and revised the manuscript.

B.J.B. contributed to study conceptualization, statistical analysis, interpretation of results, and manuscript revision.

A.L.L., J.T.J., J.D.B., N.K., A.F., C.L., K.C.C., and S.S. contributed to data acquisition and manuscript revision.

A.T.P. contributed to data acquisition, study conceptualization, statistical analysis, interpretation of results, and manuscript revision.

D.T.L. contributed to study conceptualization, statistical analysis, interpretation of results, and manuscript revision.

All authors approved the final manuscript and agreed to be accountable for all aspects of the work.

## Tables and Figures

**Supplemental Table 1.** Viral Gastroenteritis Distribution Across Age Groups (IMPACT)

| Age Group | Patients (n=337) |
| --- | --- |
| <2 years | 193 (57.27%) |
| ≥2 to <5 years | 71 (21.07%) |
| ≥5 to <12 years | 54 (16.02%) |
| ≥12 years | 19 (5.64%) |

**Supplemental Table 2.** Mixed Effects Logistic Regression of Viral vs. Non-Viral Gastroenteritis - Expanded Model (IMPACT)

| Term | OR | 95% CI | p-value |
| --- | --- | --- | --- |
| Intercept | 1.00 | 0.81 to 1.23 | <0.001 |
| Age $\geq 2$ to <5 years | 0.54 | 0.37 to 0.78 | <0.001 |
| Age $\geq 5$ to <12 years | 0.39 | 0.26 to 0.57 | <0.001 |
| Age $\geq 12$ years | 0.24 | 0.14 to 0.42 | <0.001 |
| Weighted Temperature | 0.61 | 0.50 to 0.76 | <0.001 |
| Weighted Precipitation | 1.05 | 0.88 to 1.24 | 0.611 |
| Weighted Humidity | 0.80 | 0.66 to 0.97 | 0.026 |
| Weighted Carbon Monoxide | 1.29 | 1.06 to 1.58 | 0.012 |
| Weighted Lead | 0.89 | 0.72 to 1.10 | 0.288 |
| Weighted Nitrogen Dioxide | 0.92 | 0.70 to 1.20 | 0.516 |
| Weighted Ozone | 1.12 | 0.89 to 1.42 | 0.331 |

Note: All environmental exposures were standardized (mean = 0, SD = 1). Models included site-level random intercepts. Children <2 years served as the reference age group.

**Supplemental Figure 1.**
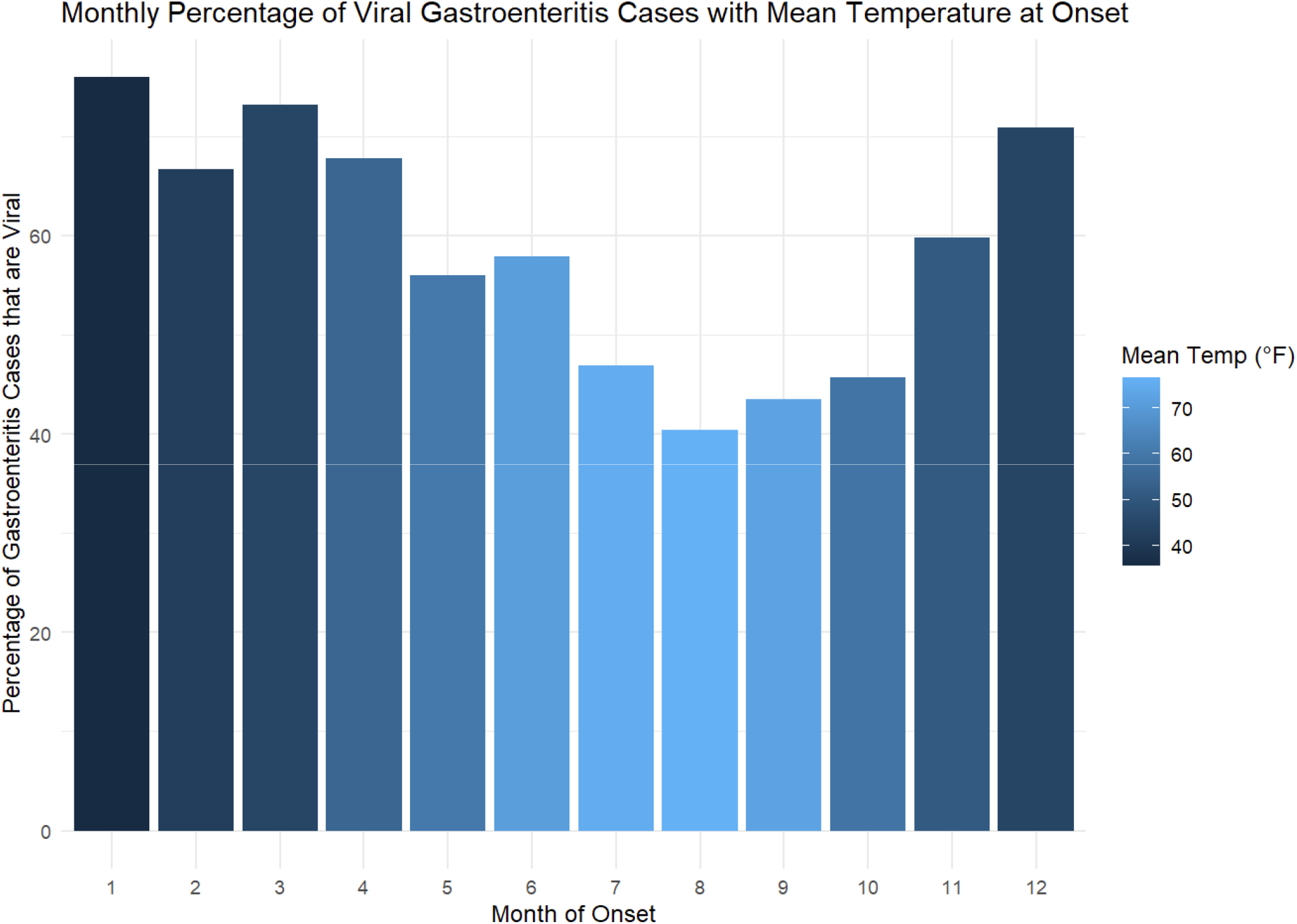
Proportion of Viral Gastroenteritis Cases by Month (IMPACT). The monthly proportion of viral IGE cases was calculated by dividing the number of viral cases by the total number of gastroenteritis cases with identified stool pathogen for each calendar month. Bars represent the percentage of cases classified as viral by month of symptom onset. Bar color indicates the mean ambient temperature (°F) during the 14 days preceding symptom onset, averaged across all cases within each month. Months with missing temperature or etiologic data were excluded

**Supplemental Figure 2.**
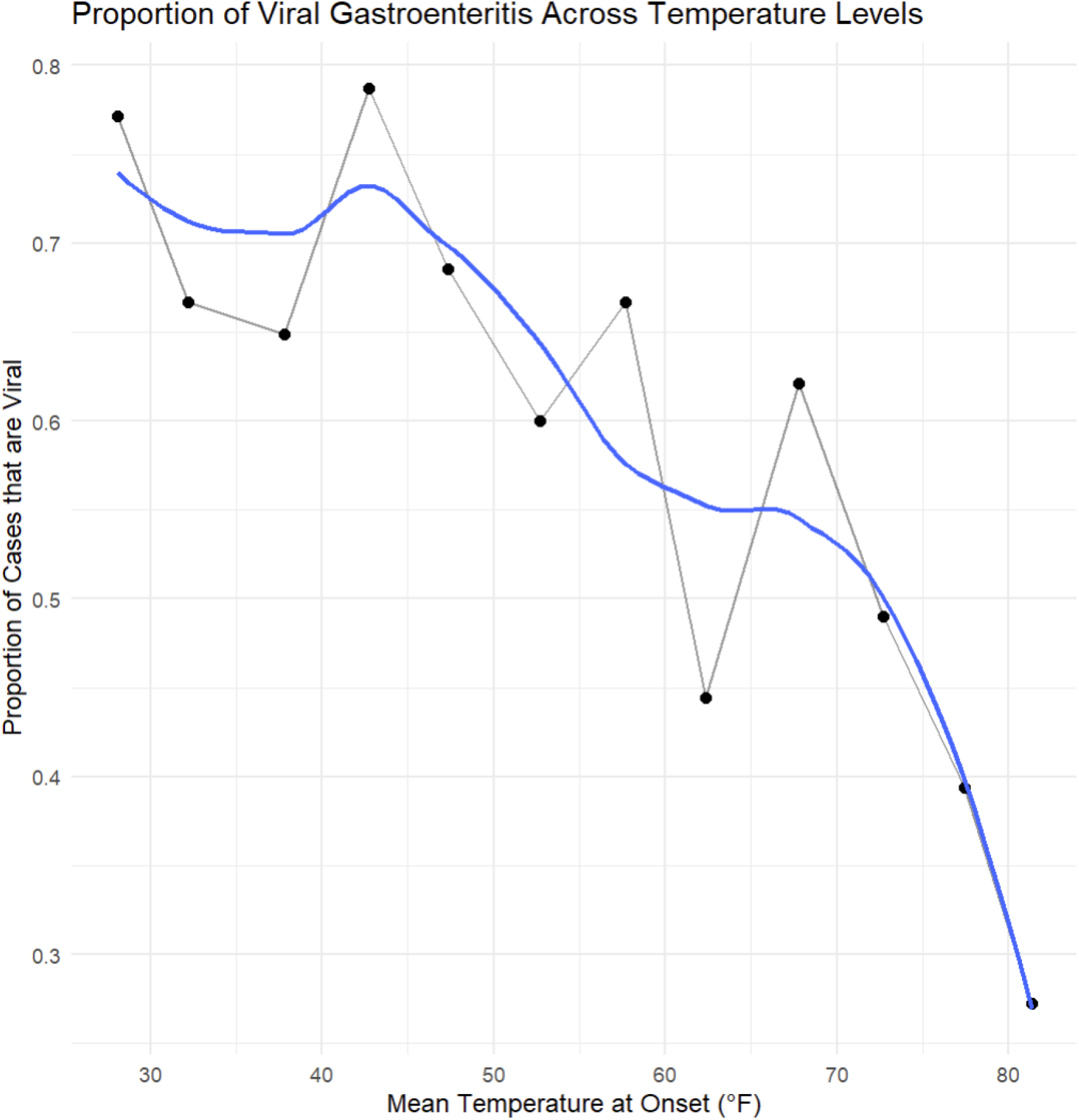
Proportion of Viral Gastroenteritis Cases by Temperature (IMPACT) Mean ambient temperature during the 14 days preceding symptom onset was grouped into 5°F bins. For each temperature bin, the proportion of viral IGE cases was calculated as the number of viral cases divided by the total number of gastroenteritis cases with an identified viral status within that bin. Points represent observed proportions, and bins containing fewer than 10 cases were excluded. The solid curve represents a locally estimated scatterplot smoothing (LOESS) fit (span = 0.6) to illustrate the overall relationship between temperature and viral etiology.

**Supplemental Figure 3.**
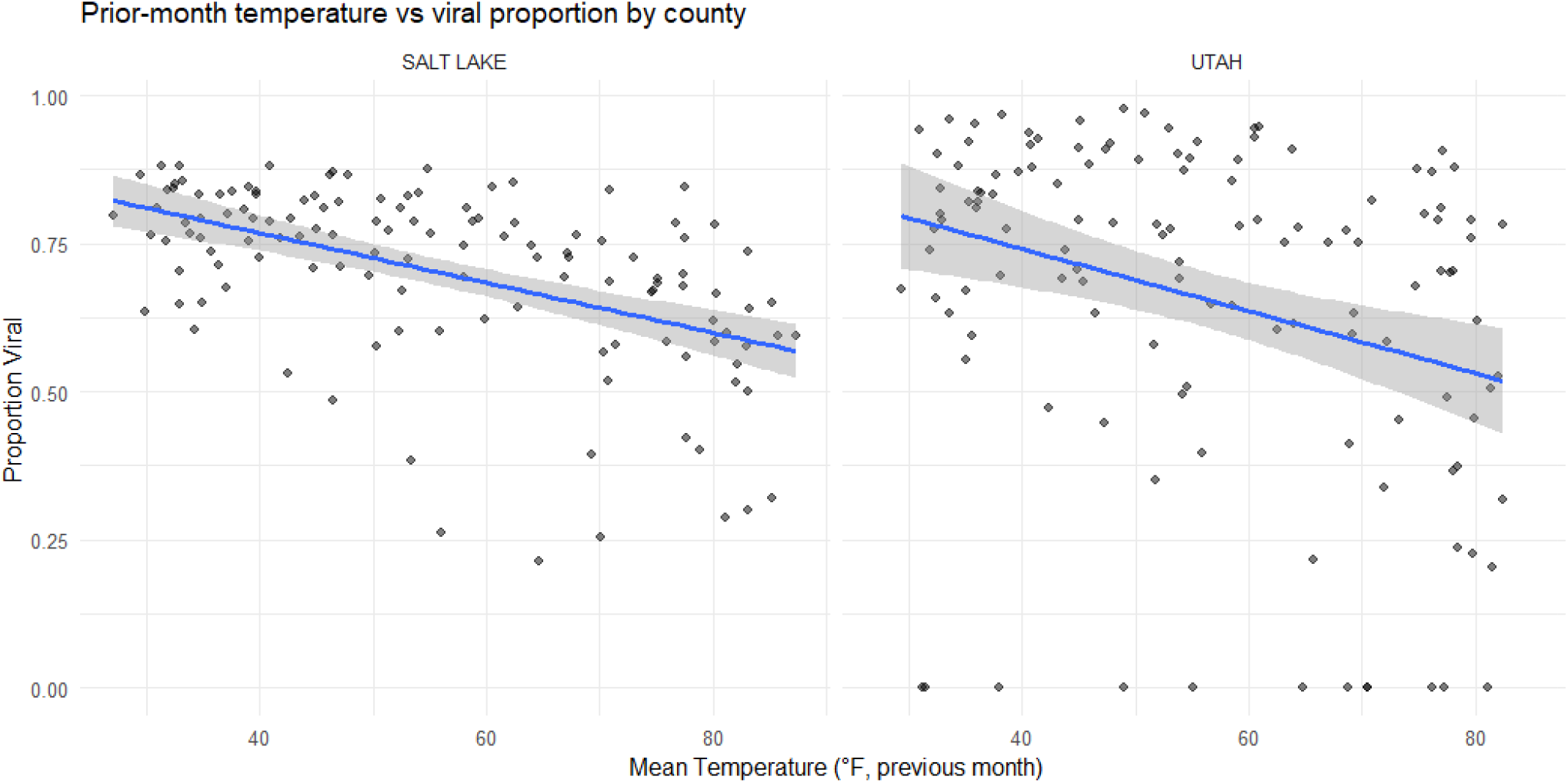
Prior Month Temperature vs. Proportion of Viral Pathogens - County-Level (GermWatch) Scatterplots depict the association between mean ambient temperature during the prior month and the monthly proportion of viral pathogen detections for Salt Lake County and Utah County separately. Monthly viral proportions were calculated as the number of viral pathogen detections divided by the total number of identified pathogens for each county-month. Solid lines represent county-specific linear regression fits with 95% confidence intervals (shaded), demonstrating consistent inverse associations between temperature and viral pathogen proportion across counties.

**Supplemental Figure 4.**
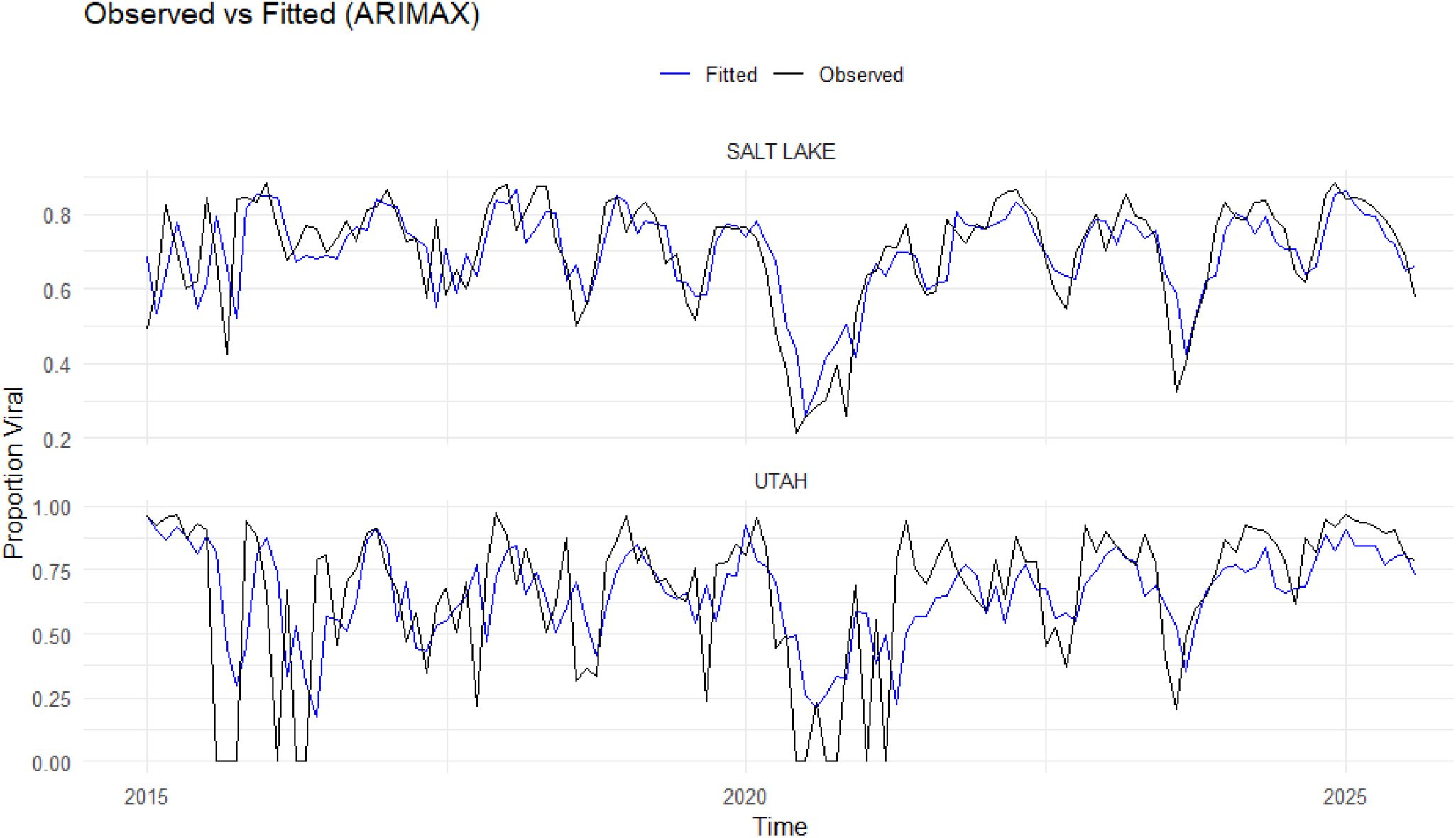
Observed and Fitted Monthly Proportions of Viral Pathogens (2015– 2025) from ARIMAX models for Salt Lake County and Utah County (GermWatch) Observed monthly proportions of viral pathogen detections (black lines) are shown alongside fitted values (blue lines) generated from autoregressive integrated moving average models with exogenous temperature inputs (ARIMAX). Monthly proportions were defined as the number of viral pathogen detections divided by the total number of identified pathogens. Models incorporated prior-month mean temperature as an external regressor to account for seasonal weather patterns. Separate models were fit for Salt Lake County and Utah County. Agreement between observed and fitted values demonstrates the ability of temperature-informed time-series models to capture seasonal variation in viral pathogen detection.

**Supplemental Figure 5.**
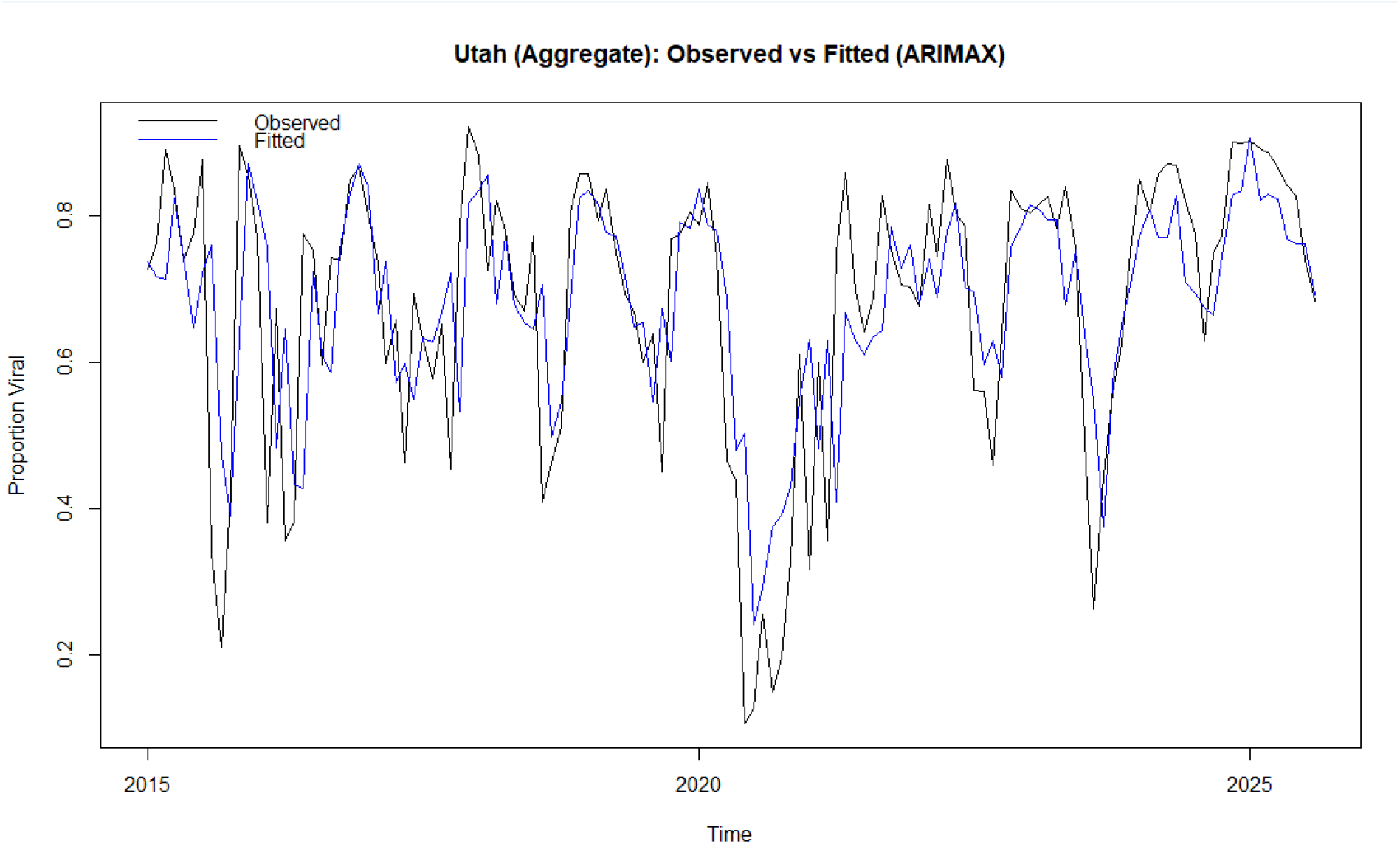
Observed and Fitted Monthly Proportions of Viral Pathogens (2015– 2025) from ARIMAX models for Salt Lake and Utah Counties (GermWatch) Observed (black line) and ARIMAX-fitted (blue line) monthly proportions of viral pathogen detections are shown for statewide aggregated GermWatch data. Monthly viral proportions were modeled using autoregressive integrated moving average models with prior-month mean temperature included as an exogenous predictor. The fitted series illustrates the contribution of temperature to explaining seasonal variation in viral pathogen detection at the population level.

